# Elevated COVID-19 Case Rates of Government Employees, District of Columbia, 2020-22

**DOI:** 10.1101/2022.01.18.22269410

**Authors:** Xinyi Hua, Jingjing Yin, Isaac C. H. Fung

**Author notes:** Corresponding author: Isaac C. H. Fung, PhD, Department of Biostatistics, Epidemiology and Environmental Health Sciences, Jiann-Ping Hsu College of Public Health, PO Box 7989, Georgia Southern University, Statesboro, GA 30460, USA. **Funding:** We do not receive any external funding to conduct this study. **Ethics:** The Georgia Southern University Institutional Review Board made a non-human subject determination for this project (H20364) under the G8 exemption category according to the Code of Federal Regulations Title 45 Part 46.

## Abstract

The rate ratios of COVID-19 cases among 5 District of Columbia departments’ personnel with reference to D.C. residents were found greater than one from March 2020 to early January 2022, suggesting higher case rates for emergency responders and frontline personnel than for general population. Vaccination is highly recommended.

**Article summary line:** Government employees from five D.C. departments, especially fire fighters and police officers, experienced higher COVID-19 case rates than D.C. residents.

---

Due to high frequency of human contacts, a higher rate of Coronavirus Disease 2019 (COVID-19) cases may be observed among first responders and essential workers than in the general population (1). Here, we estimated the rate ratios (RR) of COVID-19 cases among personnel from seven District of Columbia (D.C.) departments with reference to D.C. residents.

Downloaded from the D.C. COVID-19 surveillance repository (2), the dataset contained the number of personnel tested positive by reported date. Seven departments were selected for analysis: Fire and Emergency Medical Services Department (FEMS), Metropolitan Police Department (MPD), Department of Corrections (DOC), Department of Youth Rehabilitation Services (DYRS), the Office of Unified Communications (OUC), the Child and Family Services Agency (CFSA), and Department of Motor Vehicle (DMV). We used the full-time equivalent hours reported to estimate the employee number in each department in 2020 and 2021 (3).

With the reported case number as the dependent variable, we applied Poisson regression models with person-day as the offset to calculate the COVID-19 case RR by department, using D.C. residents as the reference. We assessed the RR over the entire study period (3/7/2020-1/3/2022), and in four distinct time periods: before COVID-19 vaccination (3/7/2020-12/14/2020), until 50% of D.C. residents were fully vaccinated (12/15/2020-6/14/2021), before 5-to-11-year-olds were eligible for COVID-19 vaccination (6/15/2021-11/2/2021), and when vaccine was available to 5-to-11-year-olds and Omicron became the dominant variant (11/3/2021-1/3/2022) (4). Statistical analysis was performed using R 4.0.3 (R Core Team, R Foundation for Statistical Computing, Vienna, Austria). The Georgia Southern University Institutional Review Board made a non-human subject determination for this project (H20364) under the G8 exemption category according to the Code of Federal Regulations Title 45 Part 46.

Five D.C. departments had higher COVID-19 case rate than D.C. residents from March 7, 2020, through January 3, 2022 (**Figure 1, Table 1**). FEMS employees’ case rate was three times that of the general population (RR=3.37; 95% CI, 3.17, 3.57). This was followed by DOC (2.67; 95% CI, 2.45, 2.91), OUC (2.23; 95% CI, 1.87, 2.63) and MPD (2.04; 95% CI, 1.93, 2.14). Stratified by pandemic stages, the RR changed over time. For example, the RR of DOC dropped from 2.88 (95% CI, 2.41, 3.41) before vaccine roll-out to 1.76 (95% CI, 1.41, 2.17) after vaccine roll-out to 1.09 (0.76, 1.51) after 50% of D.C. residents have been fully vaccinated, and increased to 3.54 (95% CI, 3.15, 3.97) as Omicron variant became dominant. Noteworthy was the observation that after 50% of D.C. residents were fully vaccinated (3^rd^ time-period), only three departments had statistically significant elevated case rates: FEMS (2.47; 95% CI, 2.05, 2.94), OUC (1.83; 95% CI, 1.08, 2.89) and MPD (1.56; 95% CI, 1.33, 1.81). As Omicron variant spread rapidly in D.C. (4^th^ time-period), five D.C. departments had statistically significant elevated case rates: FEMS (4.20; 95% CI, 3.86, 4.56), DOC (3.54; 95% CI, 3.15, 3.97), OUC (2.54; 95% CI, 1.95, 3.25), MPD (2.05; 95% CI, 1.89, 2.22), and DYRS (1.71; 95% CI, 1.30, 2.19).

**Figure 1:**
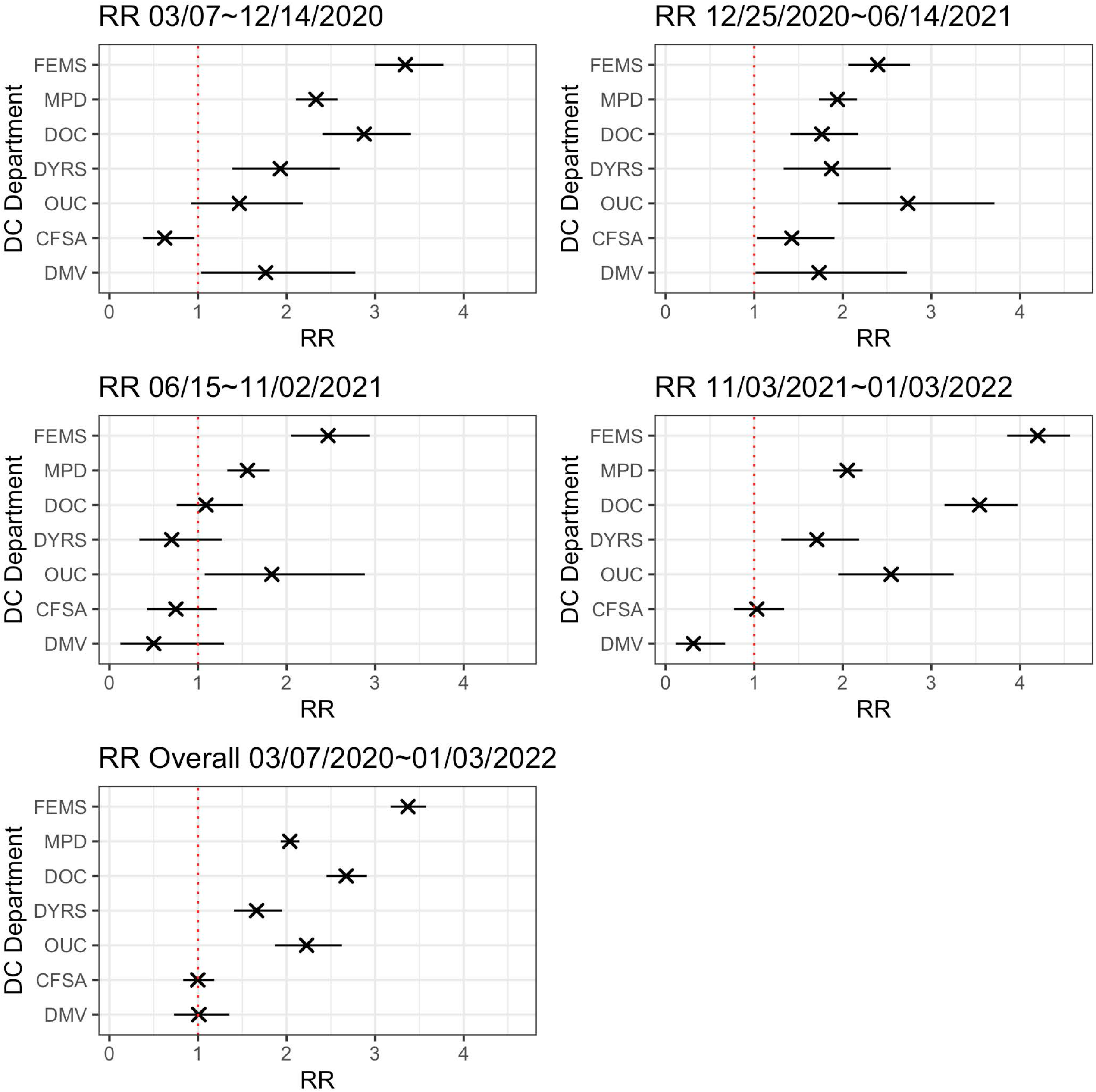
The rate ratio (cross) and 95% confidence intervals (bar) of COVID-19 case rates for seven District of Columbia departments for five selected time intervals: March 7^th^, 2020-December 14^th^, 2020, December 15^th^, 2020-June 14^th^, 2021, June 15^th^, 2021-November 2^nd^, 2021, November 3^rd^, 2021-January 3^rd^, 2022, and March 7^th^, 2020-January 3^rd^, 2022. The departments analyzed were: Fire and Emergency Medical Services Department (FEMS), Metropolitan Police Department (MPD), Department of Corrections (DOC), Department of Youth Rehabilitation Services (DYRS), the Office of Unified Communications (OUC), the Child and Family Services Agency (CFSA), and Department of Motor Vehicle (DMV).

**Table 1.** The rate ratio (RR) of COVID-19 cases of seven District of Columbia departments, from March 7^th^, 2020, to January 3^rd^, 2022.

| Departments | 3/7/2020 - 12/14/2020 |  | 12/15/2020 - 6/14/2021 |  | 6/15/2021 - 11/2/2021 |  | 11/3/2021- 1/3/2022 |  | 3/7/2020 - 1/3/2022<br>(Overall) |  |
| --- | --- | --- | --- | --- | --- | --- | --- | --- | --- | --- |
|  | RR | 95% CI | RR | 95% CI | RR | 95% CI | RR | 95% CI | RR | 95% CI |
| *Fire and Emergency Medical Services* | 3.3408 | (2.9448, 3.7710) | 2.3935 | (2.0605, 2.7609) | 2.4689 | (2.0534, 2.9375) | 4.2004 | (3.8561, 4.5649) | 3.3705 | (3.1745, 3.5744) |
| *Metropolitan Police Department* | 2.3332 | (2.1066, 2.5758) | 1.9385 | (1.7316, 2.1614) | 1.5578 | (1.3309, 1.8093) | 2.0501 | (1.8863, 2.2233) | 2.0371 | (1.9332, 2.1447) |
| *Department of Corrections* | 2.8761 | (2.4052, 3.4051) | 1.7642 | (1.4089, 2.1749) | 1.0905 | (0.7592, 1.5066) | 3.5442 | (3.1473, 3.9734) | 2.6732 | (2.4511, 2.9084) |
| *Youth Rehabilitation Services* | 1.9303 | (1.3858, 2.6018) | 1.8721 | (1.3315, 2.5425) | 0.7038 | (0.3381, 1.2689) | 1.7069 | (1.3042, 2.1851) | 1.6614 | (1.4036, 1.9490) |
| *Office of Unified Communications* | 1.4653 | (0.9244, 2.1847) | 2.7338 | (1.9444, 3.7127) | 1.8336 | (1.0752, 2.8850) | 2.5445 | (1.9489, 3.2508) | 2.2259 | (1.8686, 2.6262) |
| *Child and Family Services Agency* | 0.6249 | (0.3788, 0.9597) | 1.4248 | (1.0317, 1.9070) | 0.7504 | (0.4226, 1.2151) | 1.0297 | (0.7715, 1.3377) | 0.9974 | (0.8319, 1.1835) |
| *Department of Motor Vehicles* | 1.7649 | (1.0350, 2.7767) | 1.7313 | (1.0153, 2.7239) | 0.4997 | (0.1243, 1.2955) | 0.3134 | (0.1124, 0.6737) | 1.0095 | (0.7280, 1.3555) |

1. Residents of D.C. were used as the reference group. 2. The first COVID-19 case in D.C. was reported on March 7<sup>th</sup>, 2020. 3. The first dose of the Pfizer-BioNTech COVID-19 vaccination was administered in the United States on December 15<sup>th</sup>, 2020. 4. Fifty percent of the D.C. residents were considered fully vaccinated on June 14<sup>th</sup>, 2021. 5. The Centers for Disease Control and Prevention's Advisory Committee on Immunization Practices made an interim recommendation to use the Pfizer-BioNTech COVID-19 vaccine in teens aged 12 to 15 years on May 12<sup>th</sup>, 2021, and in children aged 5 to 11 years on November 2<sup>nd</sup>, 2021. 6. The first cases of Omicron variant in D.C. were announced on December 13<sup>th</sup>, 2021 (7).

Gradual decline in RR was observed in some departments such as DOC. This might reflect DOC effort to suppress COVID-19 transmission among its staff and residents as described elsewhere (5). However, DOC had a major surge among its personnel and residents when the Omicron variant was rapidly spreading in D.C. in the 4^th^ time period (6).

The estimated RR of FEMS and MPD were consistently greater than one, both overall and by each time period, reflecting that their employees have been on the front lines of providing support and services to the local community as the pandemic continues. Overall, personnel from five out of seven D.C. departments experienced higher COVID-19 case rates than D.C. residents.

Our study had limitations. First, the data were analyzed by report date as symptoms onset date was unavailable. Second, asymptomatically infected individuals might not get tested; case rate is an underestimate of the underlying infection rate. Third, gender and age-specific RR by occupation were unavailable as such demographic information of D.C. governmental employees was not publicly available.

Five D.C. departments had occupation-specific elevated COVID-19 case rates in 2020-22. Government employees providing service in the front lines should consider vaccination (including boosters) to protect themselves against COVID-19.

## Data Availability

This study involves only openly available human data, which can be obtained from the Government of the District of Columbia website: https://coronavirus.dc.gov/data
All results generated from our analysis are presented in the manuscript.

## First author’s bio

Ms. Xinyi Hua is a doctoral student in epidemiology at Georgia Southern University. Her research interest is in infectious disease epidemiology.

